# Implementation of risk triaging in primary healthcare facilities in Sub-Saharan Africa: A systematic review

**DOI:** 10.1101/2023.07.11.23292524

**Authors:** Mhairi Maskew, Linda Alinafe Sande, Mariet Benade, Vinolia Ntjiekelane, Nancy Scott, David Flynn, Sydney Rosen

## Abstract

**Background:** One challenge facing treatment programs for HIV and other chronic conditions in sub-Saharan Africa (SSA) is how to target interventions to optimize retention in care and other outcomes. Most efforts to target interventions have identified predictive features among high risk patients after negative outcomes have already been observed. An alternative for identifying patients at high risk of negative outcomes is “risk triaging,” or identifying vulnerable or higher risk patients before they experience an interruption in care or other negative outcome. We conducted a systematic review of the use of risk triaging tools at the primary healthcare (PHC) level in SSA.

**Methods:** We searched PubMed and other databases for publications after 1 January 2012 that reported development or implementation of risk triaging tools for PHC use in SSA. We extracted information on three outcomes: 1) characterization of the risk triaging tools; 2) tool performance metrics (sensitivity, specificity, positive and negative predictive value, area under the curve); and 3) health system effects (efficiency, acceptability, resource utilization, cost). We report outcomes for each eligible study and identify lessons for use of risk triaging.

**Results:** Of 1,876 articles identified, 28 were eligible for our review. Thirteen addressed HIV, 10 TB, 1 TB/HIV, and 4 other conditions. Approximately 60% used existing, retrospective data to identify important risk factors for an outcome and then construct a scoring system, but no implementation of these tools was reported. The remaining 40% designed a tool using existing data or experience and reported implementation results. More than half (16/28, 58%) of the tools achieved sensitivities >80%; specificity was much lower. Only one tool, the World Health Organization’s 4-symptom screen for tuberculosis, had been scaled up widely. While most studies claimed that their tools could increase the efficiency of healthcare delivery, none of the studies provided examples of tangible health system impacts.

**Conclusion:** Most of the tools identified were at least somewhat successful in identifying potential risks but uptake by health systems has been minimal. Although well-designed risk triaging tools have the potential to improve health outcomes, implementation will require commitment at the policy, operational, and funding levels.

## INTRODUCTION

One of the challenges facing treatment programs for HIV and other chronic conditions in sub-Saharan Africa is how to target interventions to maximize long-term retention in care [1–4]. Ideally, interventions aimed at improving adherence and retention should be offered to patients at higher risk of disengagement from care or poor adherence, while not adding to the burden of care or utilizing resources unnecessarily for lower risk patients who do not require additional attention. Identifying high risk patients before they experience negative outcomes, however, remains a puzzle.

Traditionally, most efforts to target interventions aimed at improving retention in care or reducing interruptions in treatment have identified high risk patients based on observed behaviour after negative events have occurred. Patients who are observed to miss clinical visits or medication refills, for example, are identified after the event and targeted for tracing, counselling, home-based care, and other services that may help them resume scheduled care [5–10]. An alternative to this post-hoc strategy for identifying high risk patients is “risk triaging,” or the process of identifying vulnerable or higher risk patients before they experience an interruption in care. Predictive models developed into risk scoring or triaging tools have a long history of use in hospitals, in high income countries, and for non-communicable conditions [11–13].

While risk triaging or risk scoring can be a straightforward approach for evaluating clinical symptoms, gauging a patient’s risk of a behaviour such as disengaging from chronic disease care is much more challenging. Some broad (and largely unmodifiable) patient characteristics, such as sex and age, have consistently been associated with higher loss to follow up rates in the literature[14,15] but these do not identify subgroups that allow for practical or efficient targeting of interventions. Recognizing this, multiple attempts have been made to identify patients who would benefit from early intervention. These include tools developed to identify patients at risk of ART default and poor viral load outcomes [16–19]; patients qualifying for same day ART initiation [20,21]; adults and children in need of HIV testing [22,23]; and patients likely to return to care after disengagement [24]Results of these tools have been mixed, in terms of both accuracy and uptake by healthcare providers. While much risk triaging is conducted in hospitals and other high-care settings, it is at primary healthcare clinics that most patients in SSA initiate and sustain ART. To our knowledge, the only example of HIV-related risk triaging commonly used at the primary healthcare clinic level in SSA is the World Health Organization’s symptom screen for tuberculosis, which triages patients with cough, fever, night sweats, or weight loss for TB diagnostic tests [25–27].

The absence of risk triaging for patients on ART precludes providers from being able to target specific types of support to specific patients who need that support. Patients who anticipate that transport fares will pose an obstacle to visit attendance, for example, could be offered different interventions from those who fear accidental disclosure of their HIV status, if these obstacles were identified in advance. As part of an effort to design differentiated models of HIV treatment delivery for patients in their first six months after antiretroviral therapy initiation, we are developing a risk triaging tool that could be used by clinical and lay providers to match retention interventions to levels and types of risks[28,29]. To lay the groundwork for this effort, we conducted a systematic review of the use of risk triaging tools at the primary healthcare level in SSA. We asked two main questions: 1) Can risk triaging tools accurately stratify patients into higher and lower risk groups in primary healthcare settings? and 2) How has risk triaging been implemented in primary healthcare facilities in SSA?

## METHODS

### Search strategy and study selection

For this review, we defined risk triaging or a risk scoring “tool” as any score, system, test, or algorithm that was designed to be used at point of care to differentiate participants into risk categories based on their individual risk of the pre-defined study outcome. We note that we were seeking tools that predict risk of an existing or future outcome, not diagnostic tools that confirm the presence of an existing condition. (For example, we regarded the World Health Organization symptom screen for tuberculosis to be a risk triaging tool, and thus included it in our review, while TB diagnostic tests were excluded.) Risk may pertain to an existing condition such as TB, in which case risk triaging selects a subset of all potential patients for further diagnostic investigation. This category of risk triaging usually happens when the diagnostic process is too expensive, invasive, or otherwise undesirable to offer to all patients. Alternatively, risk may pertain to a condition that does not exist yet but is more likely in some individuals than in others, such as future disengagement from treatment. Risks in this category may be amenable to intervention before the condition occurs. (We also acknowledge that the use of the term “risk” has been criticized for appearing to place blame on or stigmatize those labeled as high risk. Since “risk” remains a standard term in the literature, however, we have chosen to continue to use it here.)

We searched peer-reviewed publications and conference abstracts that reported development and/or implementation of risk triaging tools published after 1 January 2012. We limited the review to studies reporting entirely or primarily outcomes from sub-Saharan Africa and restricted our search to tools designed for or implemented in primary health care settings; any tools that required hospital in-patient admissions or required use of specialized laboratory services not typically available in outpatient settings were excluded. We considered community-based and other off-site service delivery models as outpatient primary health services and included these. Mathematical models were included if they served as a data source or as part of the process for developing a risk triaging tool but were not defined as tools themselves, and thus studies reporting only the development of mathematical models were excluded. Studies that reported purely qualitative data or were limited to theoretical concepts were excluded. A full list of inclusion and exclusion criteria and our search strategy are provided in S1 Table. The protocol for this systematic review is included as S1 Text and was registered on PROSPERO (CRD42022328209) and followed PRISMA guidelines.

The database search was conducted on 24 May 2022 and updated on 25 July 2022. We searched the following databases: NLM’s PubMed, Wiley’s Cochrane Library, the Cumulative Index of Nursing and Allied Health Literature (CINAHL) via Ebsco; EMBASE via Elsevier; Clarivate’s Web of Science (specifically the Web of Science Core Collection); and PsycINFO via Ebsco. Composite search strings were developed using keywords according to the Cochrane Collaboration guidelines’ PICO model criteria [30]. We then utilized the PICO components (populations, interventions, comparators, outcomes) to develop the composite search terms. The full list of search terms for each database searched is provided in S2 Table.

After the search was completed and results deduplicated, two authors (LS and VN) conducted an initial, blinded, independent screening of article titles and abstracts for eligibility. Conflicts were reviewed after unblinding and resolved between the screening authors and a third author (MM). Full texts of potentially eligible articles were retrieved, imported to a reference management program, and evaluated for inclusion in the review. Reasons for exclusion after full-text review were recorded. Using a snowball approach, we also manually searched reference lists of articles considered relevant to identify other sources for potential inclusion. The search results are summarized in accordance with the PRISMA-P reporting protocol (S3 Table).

### Outcomes

For purposes of this review, we were interested in both the tool’s statistical performance metrics and the extent to which the tool was taken up for routine practice, including the method of implementation, its usefulness in practical stratification of patient populations, and effects on the healthcare system. The primary outcomes for this review were thus threefold: 1) Characterization of the risk triaging tools; 2) tool performance metrics (sensitivity, specificity, positive and negative predictive value (PPV and NPV), and area under the curve); and 3) health system considerations including efficiency of clinic operations, patient waiting times, provider acceptability, resource utilization, cost, and scalability.

### Data extraction and analysis

We developed a standardized form to extract key information from each eligible article, including (1) article details; (2) patient population; (3) description of the risk triaging tool; (4) risk triaging tool performance metrics and (5) effect of the risk triaging tool implementation on the health system. After reporting this information descriptively, we plotted performance metrics to present point estimates for sensitivity and false positive rates (1-specificity). Where available, negative predictive value (NPV), positive predictive value (PPV), and area under the curve (AUC) were also plotted as individual points.

Because of the tremendous heterogeneity in the sources included in this review, which varied by condition addressed, risk triaging approach, types of tools, specific outcomes predicted by the tools, population studied, and many other parameters, we did not attempt to conduct any pooled analyses or generate aggregate values to represent the full set of sources, which we believe would present misleading summary values. We instead present the results individually, with sufficient information about each tool to allow results to be compared as appropriate to each reader’s needs.

### Methodological quality assessment

Risk of bias for each study was assessed using the Joanna Briggs Institute (JBI) critical appraisal checklist for diagnostic test accuracy studies [31]. The JBI checklist asks a set of 10 closed ended questions for every study included to evaluate risk of bias in the design, conduct and analysis in the studies meeting the inclusion criteria. A “yes” response indicates the criterion for risk of bias is low or not present while “no” responses indicating risk of bias is present. We assigned a score of 1 point to each “No” response and calculated a bias risk percentage score using the total number of questions for each study for which we were able to assess a response as the denominator. We then set the following thresholds for risk assessment: 1) score of 20 percentage points or less indicated low risk of bias; 2) score of 20-50 percentage points indicated moderate risk of bias and; 3) score of more than 50 percentage points indicated high risk of bias.

## RESULTS

### Search results

Our search strategy yielded a total of 1,876 articles from the six databases searched (Figure 1). We removed 107 articles duplicated across databases and screened abstracts and titles from 1,769 articles. Of these, 42 articles met the inclusion criteria. Of 1,708 articles that did not meet the inclusion criteria, some were excluded for multiple reasons, but having the wrong study design (n=1,201), no evidence of a risk triaging tool being implemented in the study (n=989), or involving the wrong study population (n= 745) were the most frequent reasons for article exclusion. From abstracts, we also identified 19 published systematic reviews to include in the snowballing process. We reviewed the reference lists of these articles and identified a further 40 articles meeting our inclusion criteria, for a total of 82 articles extracted for full text review. We excluded 54 of these; half of these exclusions were due to either study locations outside sub-Saharan Africa (n=14) or not in a primary health setting (n=12).

**Figure 1:**
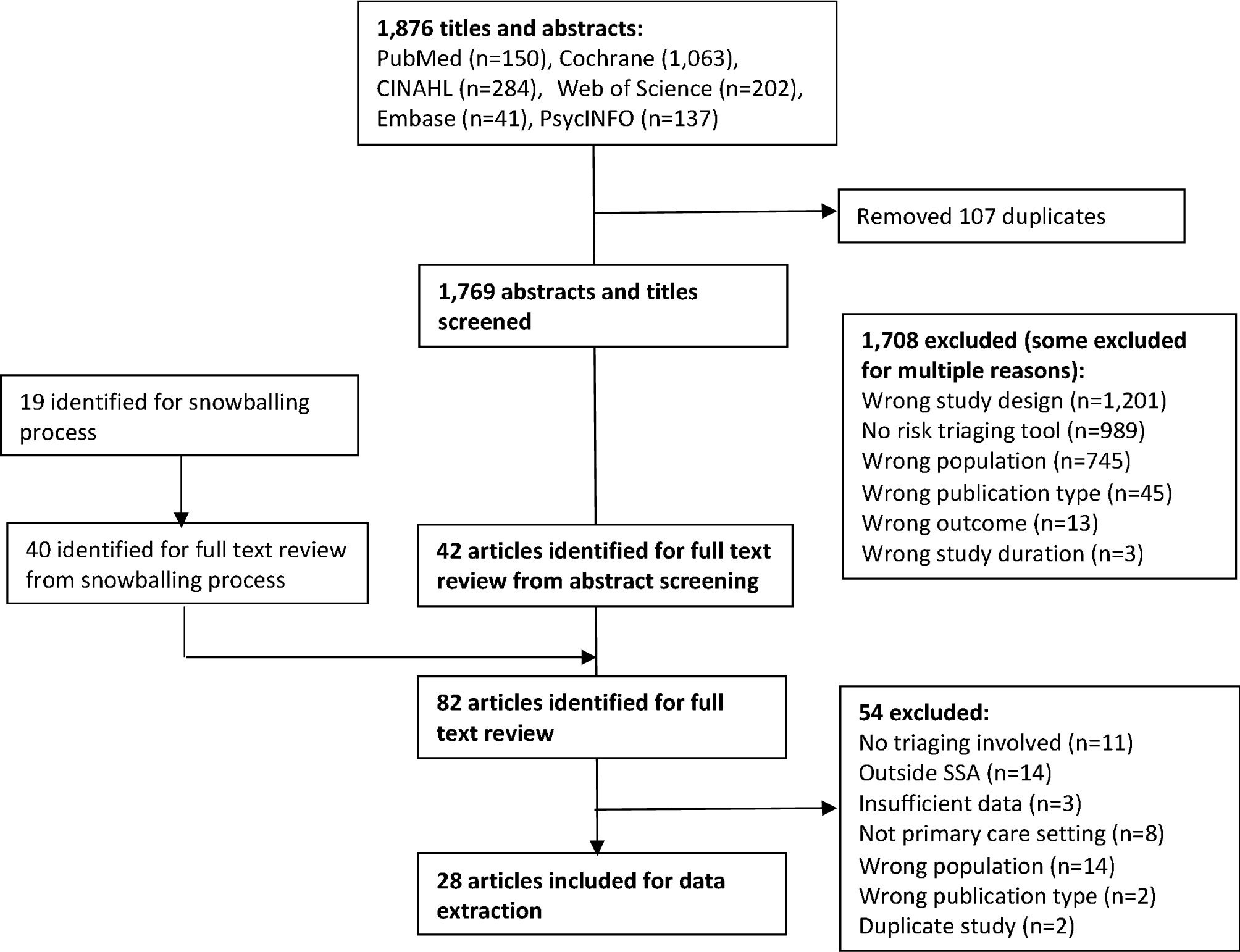
PRISMA flow chart of article screening process.

The final data set included 28 articles (Table 1) that reported on risk triaging tools from 14 observational cohorts, 6 clinical trials, and 8 other study designs, with data collected and analysed between 2004 and 2020. The most frequent implementation countries were South Africa (n=12), Kenya (n=7), and Uganda (n=6); 8 were multi-country studies. Risk triaging tools were implemented in a diverse range of facility settings within the context of primary health care, including health facility-based outpatient clinics, community-based sites, and dedicated research facilities

**Table 1:**
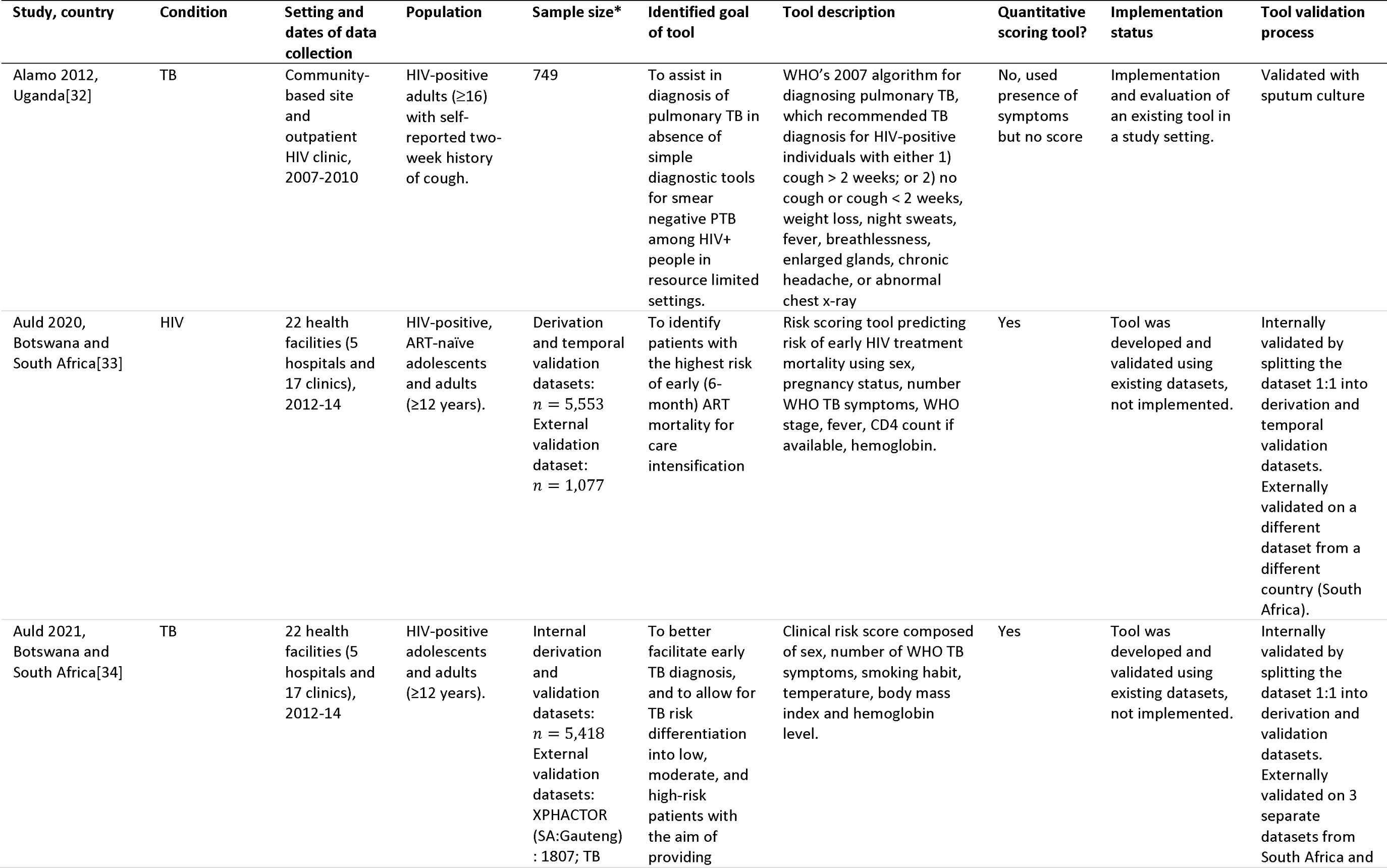
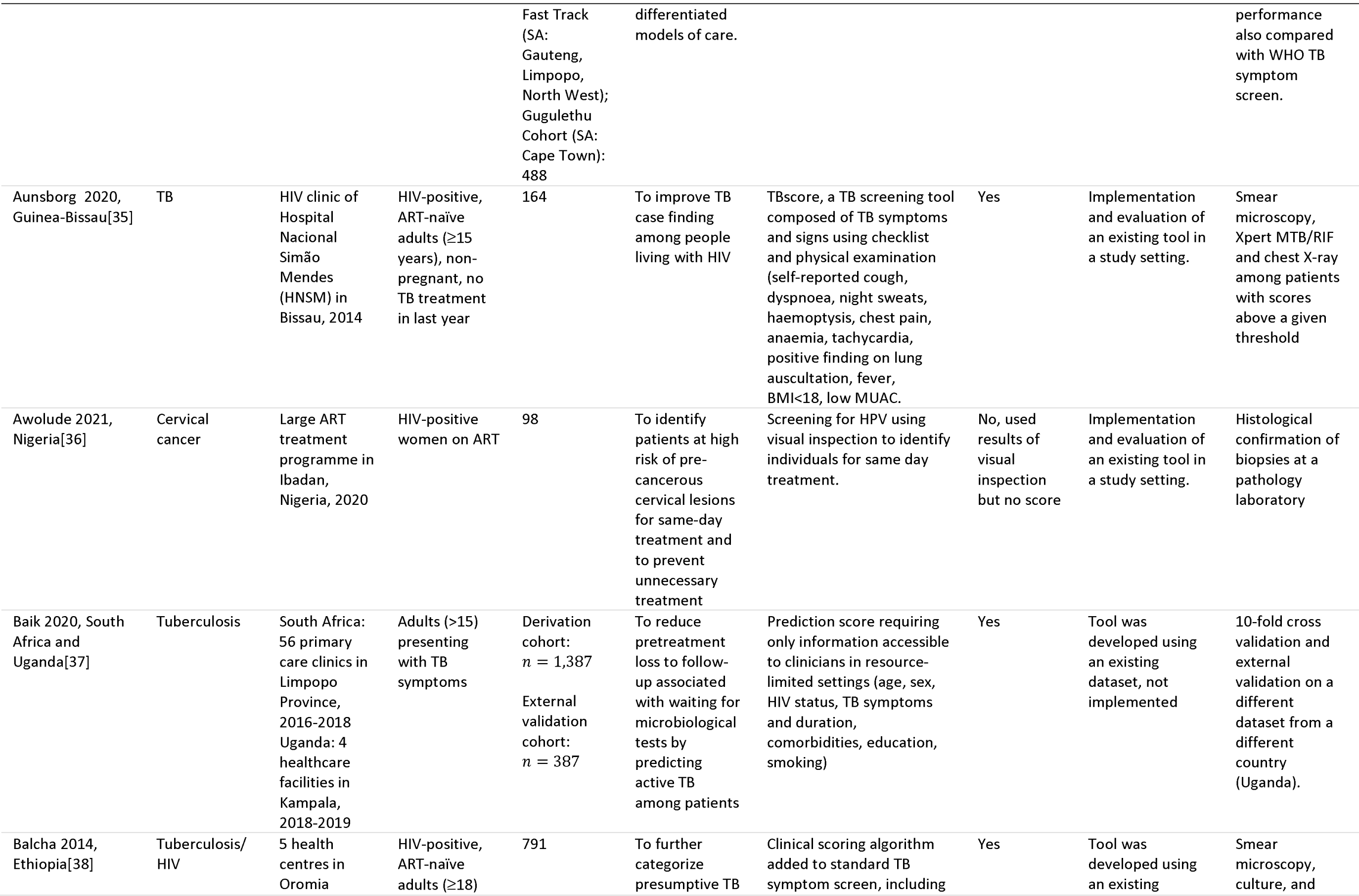
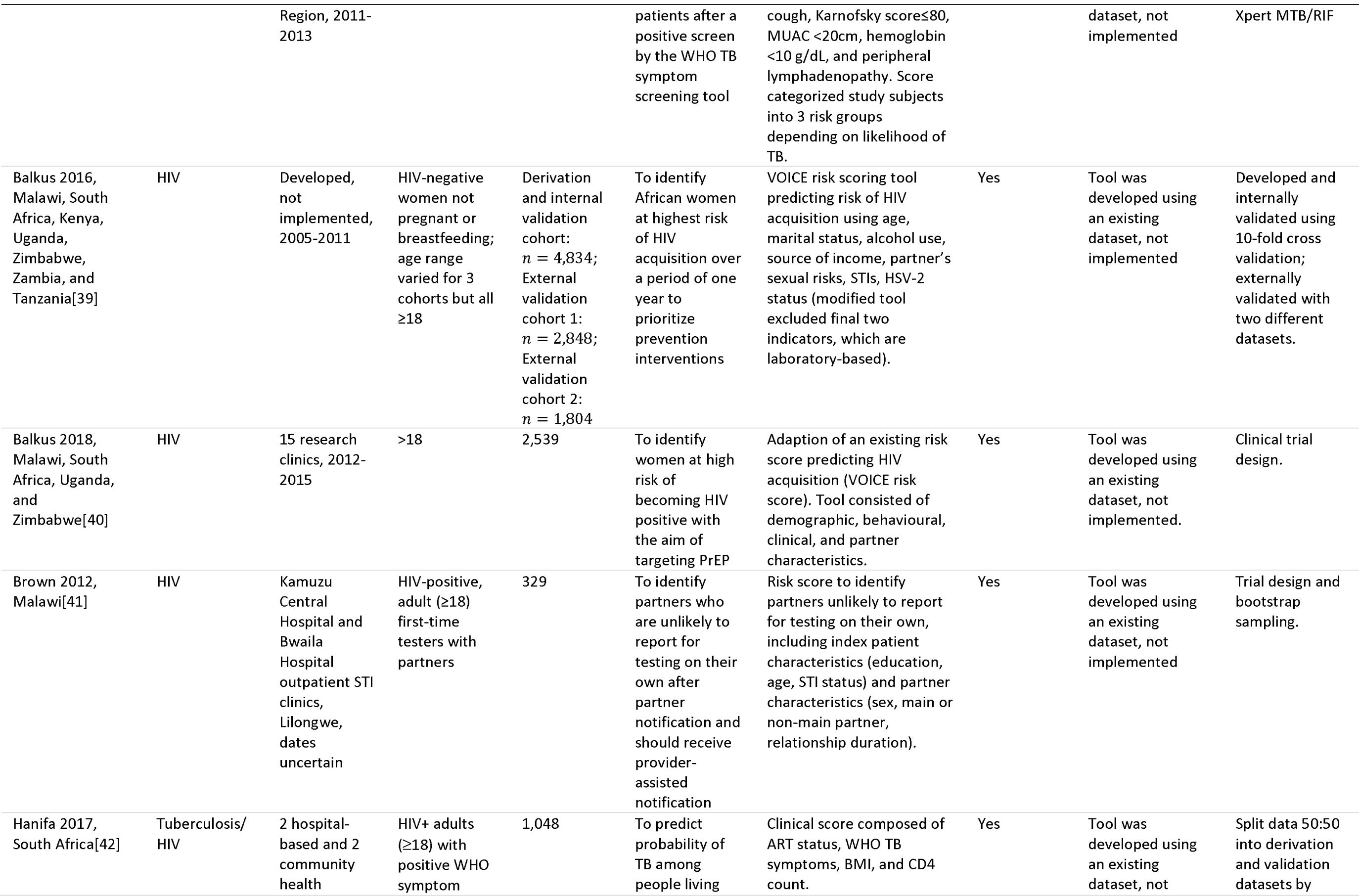
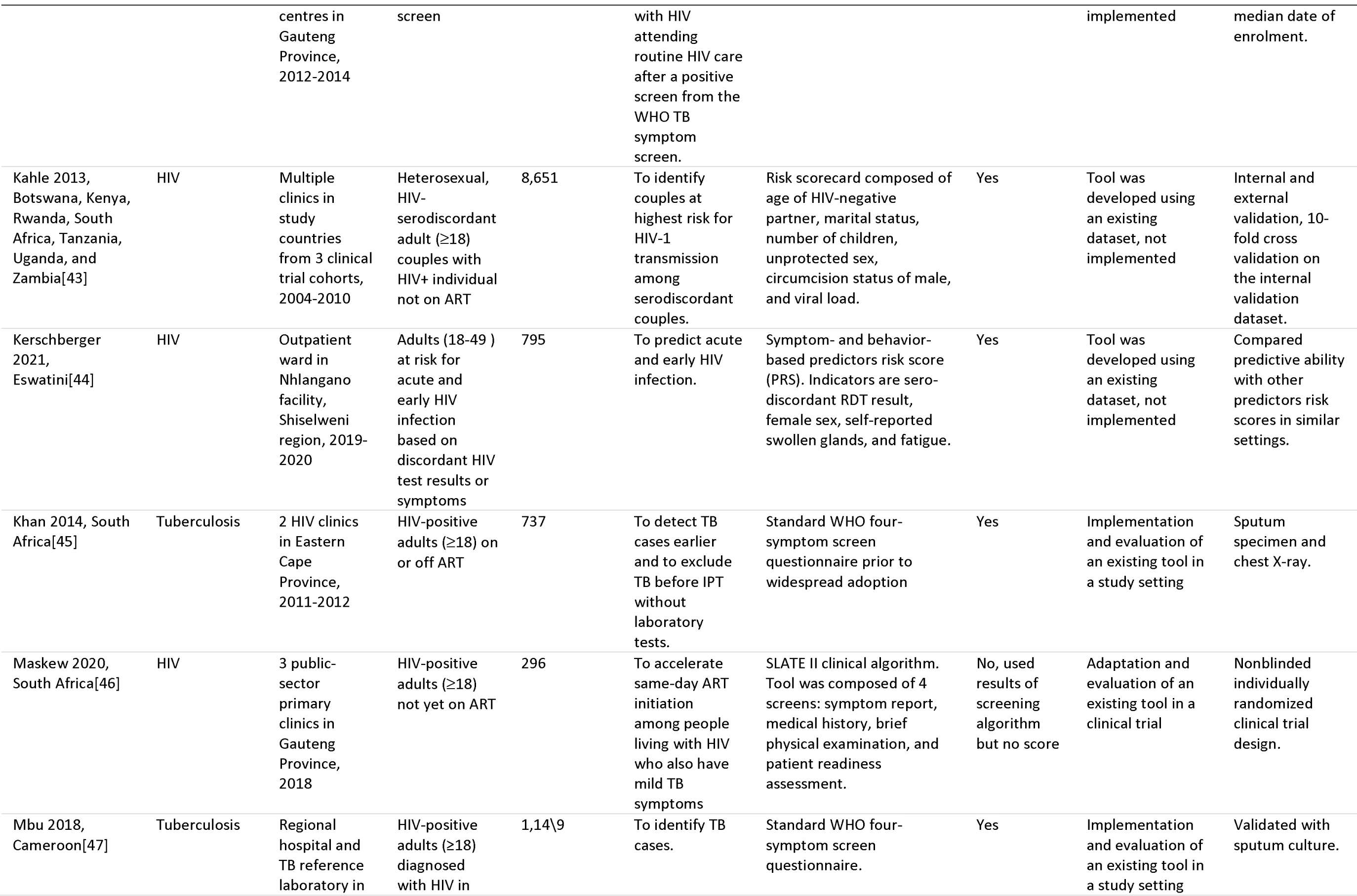
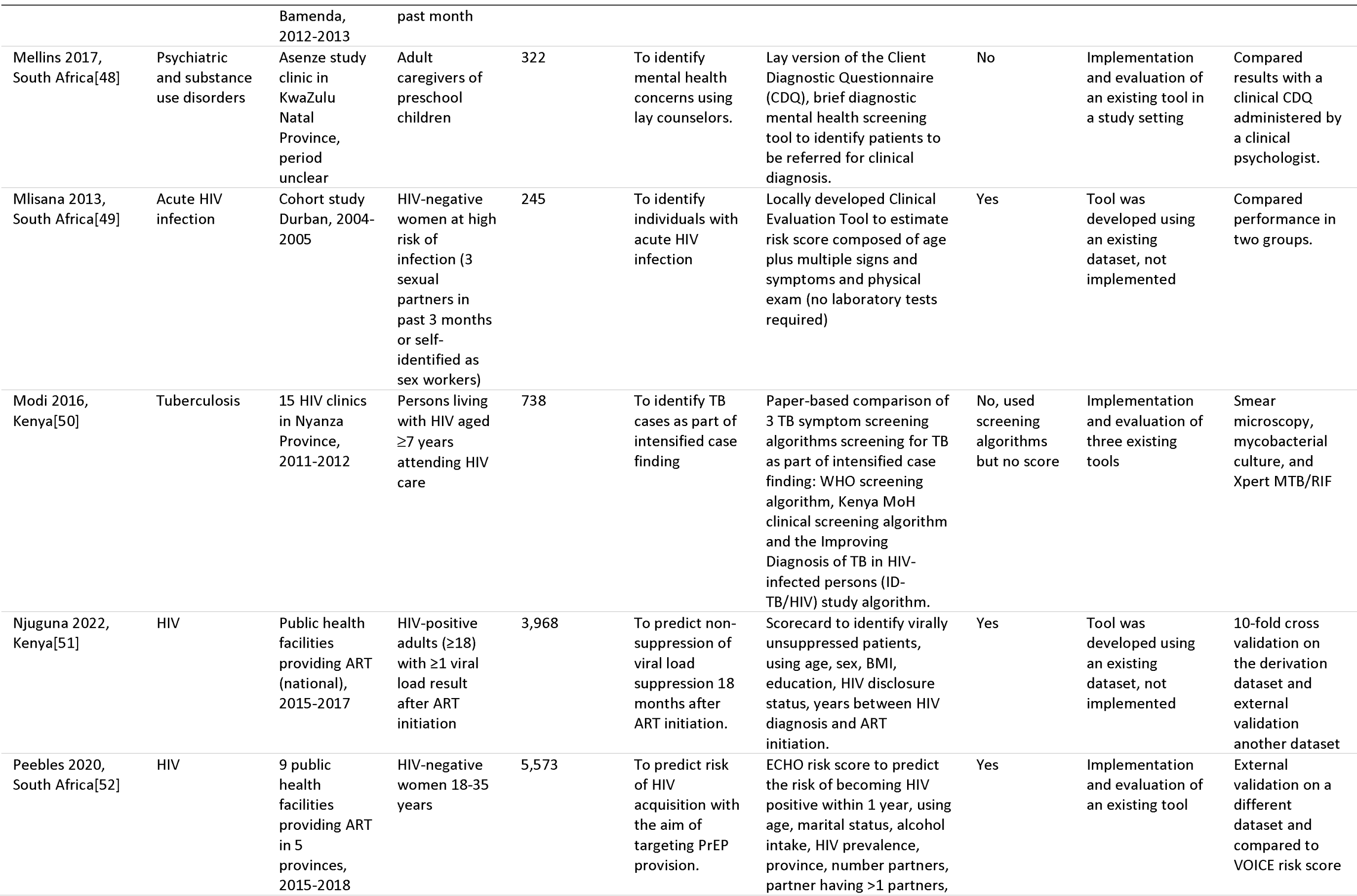
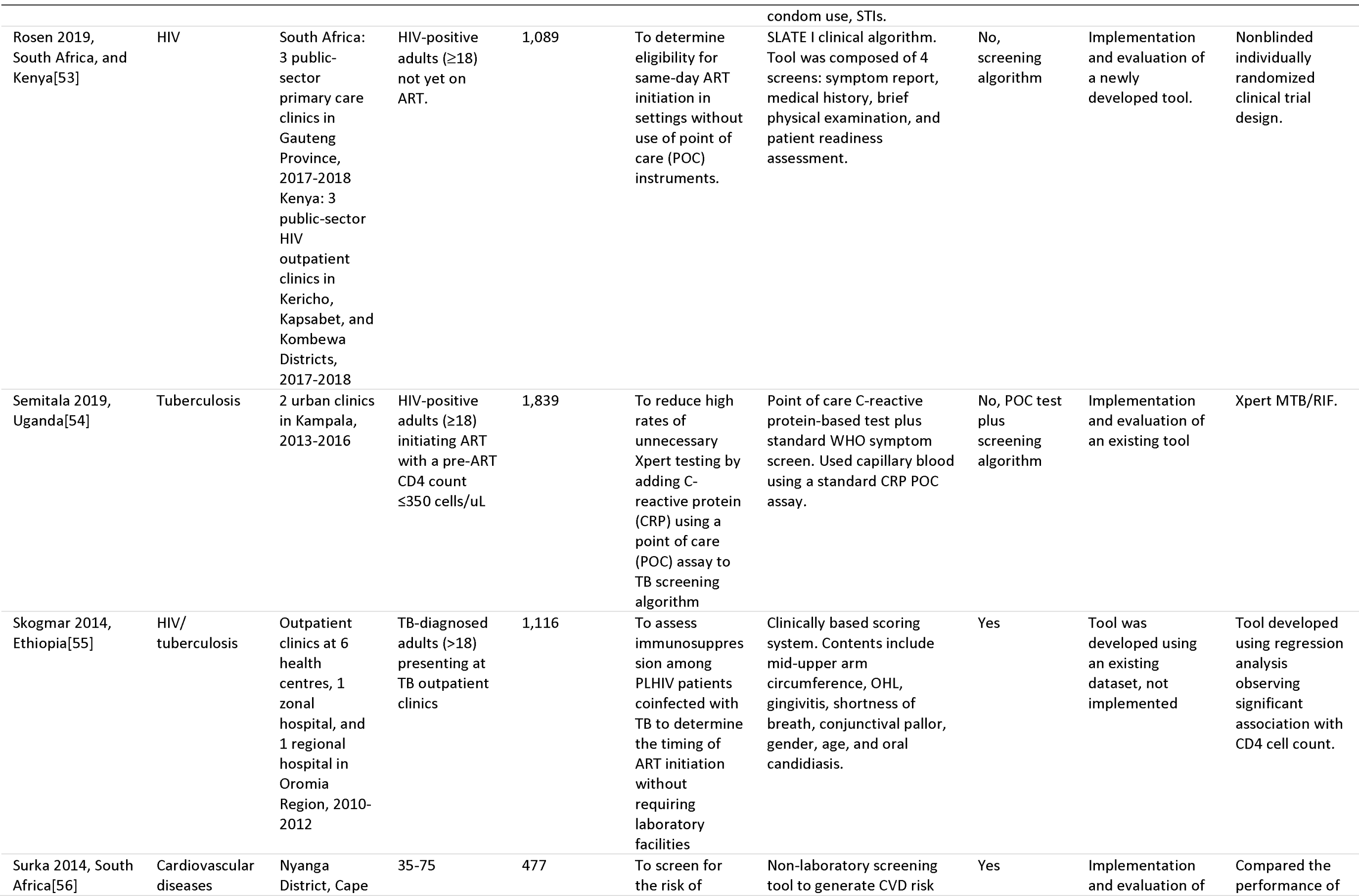
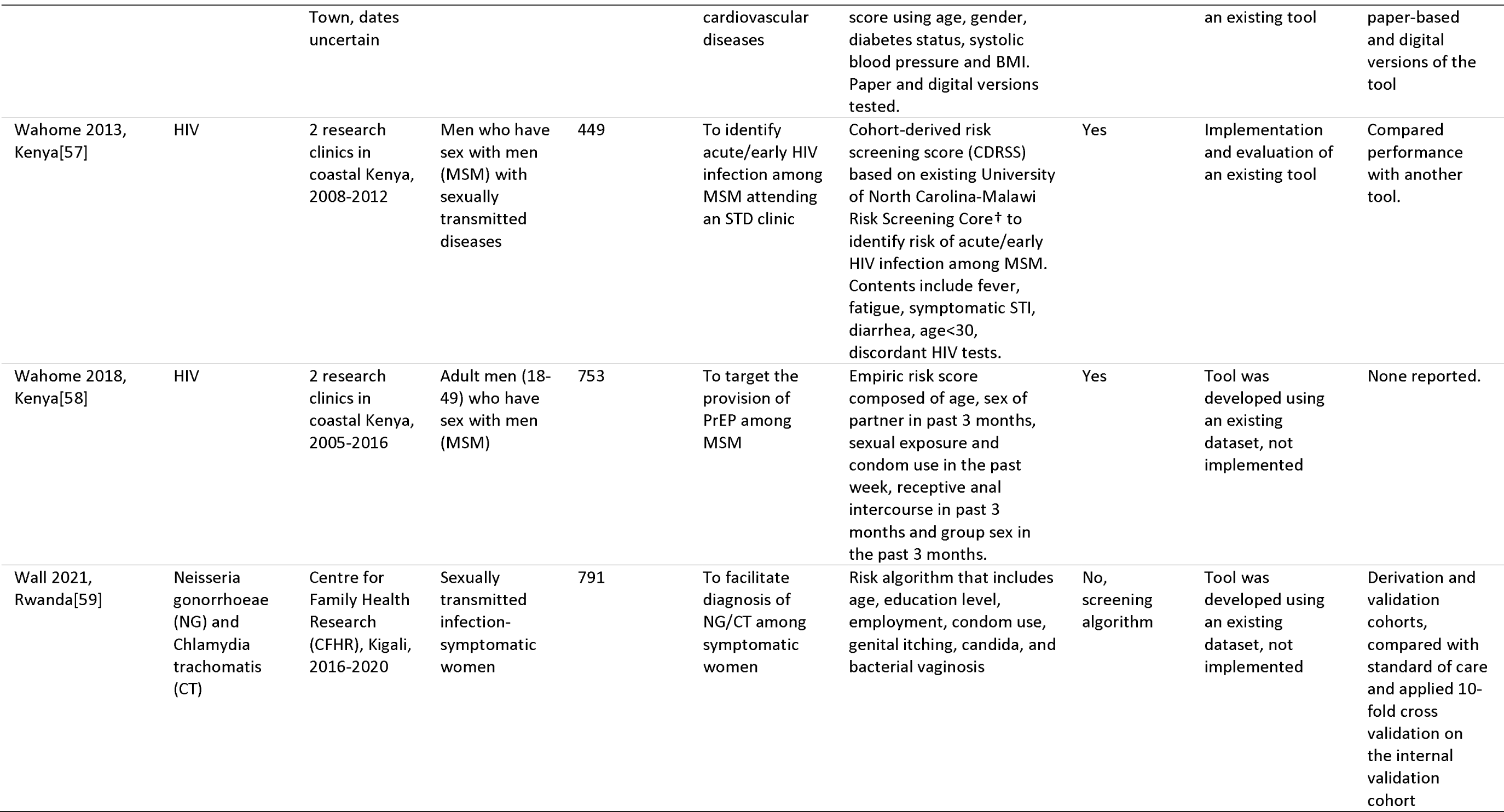
Description of risk scoring and triaging tools.

### Characterization of risk triaging tools evaluated

Table 1 and Figure 2 describe the risk triaging tools identified in our search. Of the 28 studies, 13 addressed HIV, 10 TB, 1 TB/HIV, and 4 other conditions. In total, 24 (86%) of the triaging tools generated risk scores, while the remaining 4 utilized clinical algorithms, symptom screens, or clinical checks and diagnostics. The risk score approach was most frequently applied to HIV (n=13) and TB (n=8), while the other triaging approaches were applied to HIV and TB combined, sexually transmitted diseases, cancer, and mental health conditions.

**Figure 2.**
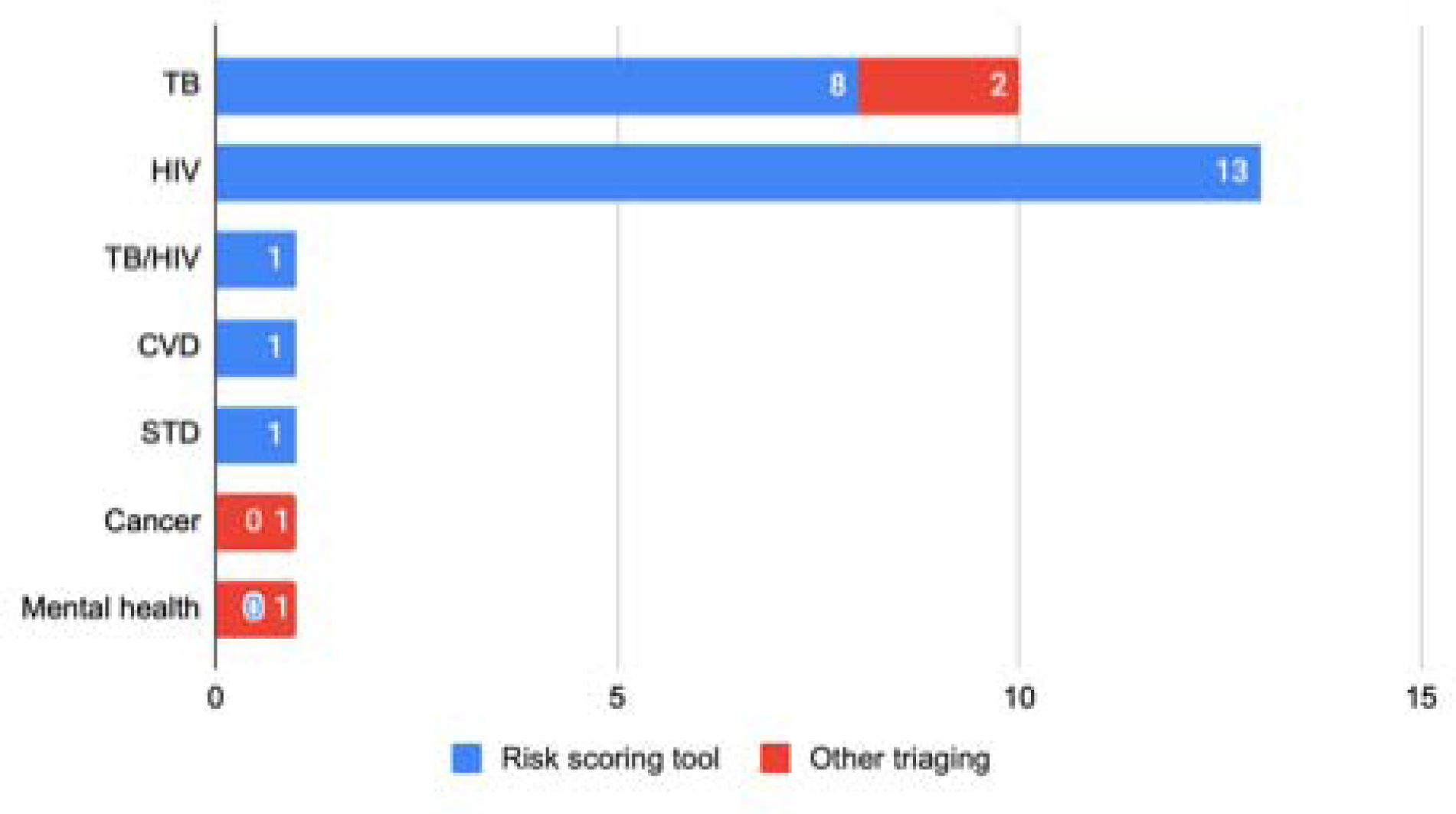
Characterization of triaging tools stratified by condition for which the tool was developed (n=28)

The studies included in the review fell into two methodological categories. First, many studies used existing, retrospective data from cohort studies (n=9) or clinical trials (n=8) to identify important risk factors for an outcome and then used these to construct a scoring system. The published reports for these tools show no evidence of actual administration of the tool to a patient cohort and appear not to have been implemented at all. A second approach was to design a tool in advance—presumably based on existing data and/or clinical experience—and then report a study in which the tool was implemented in a trial or observational cohort setting (n=11). Most of the studies in both categories developed new tools, but others compared an existing or adapted tool to standard of care or to one or more other tools, such as comparing WHO’s latest TB symptom screening algorithm to its previous version. The tools that were administered in implementation studies used digital and/or paper formats for data collection; most did not report the format utilized.

### Performance metrics

Performance metrics for the risk triaging tools evaluated are detailed in Table 2. Sensitivity of the applied tools varied between studies but also within studies that considered multiple thresholds were considered. More than half (n=16; 57%) achieved sensitivities >80%. As would be expected for risk triaging tools (in contrast to diagnostic tools), results for specificity were generally low. Of the 20 articles that reported specificity, 8 (40%) had a specificity <50%. Not all studies reported measures for negative and positive predictive value; those that did indicated negative predictive values ranging from 4%-99% and positive predictive values ranging from 1%-99%. Area under the curve (AUC) was also not universally reported, but for studies where this metric was available (n=16), most AUC values were >0.7, indicating that the tools correctly identified the outcome of interest in 7 or more of 10 patients.

**Table 2:**
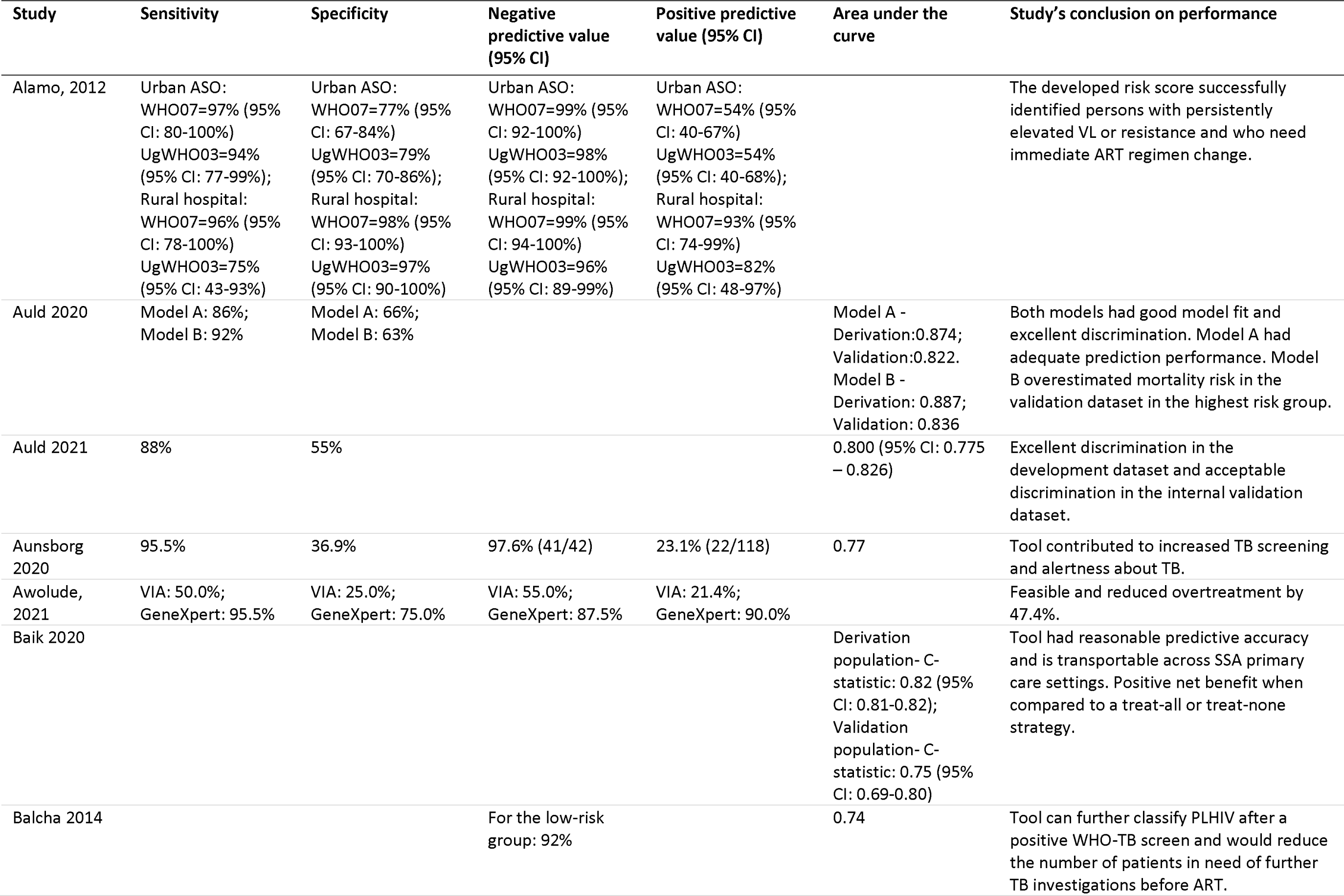
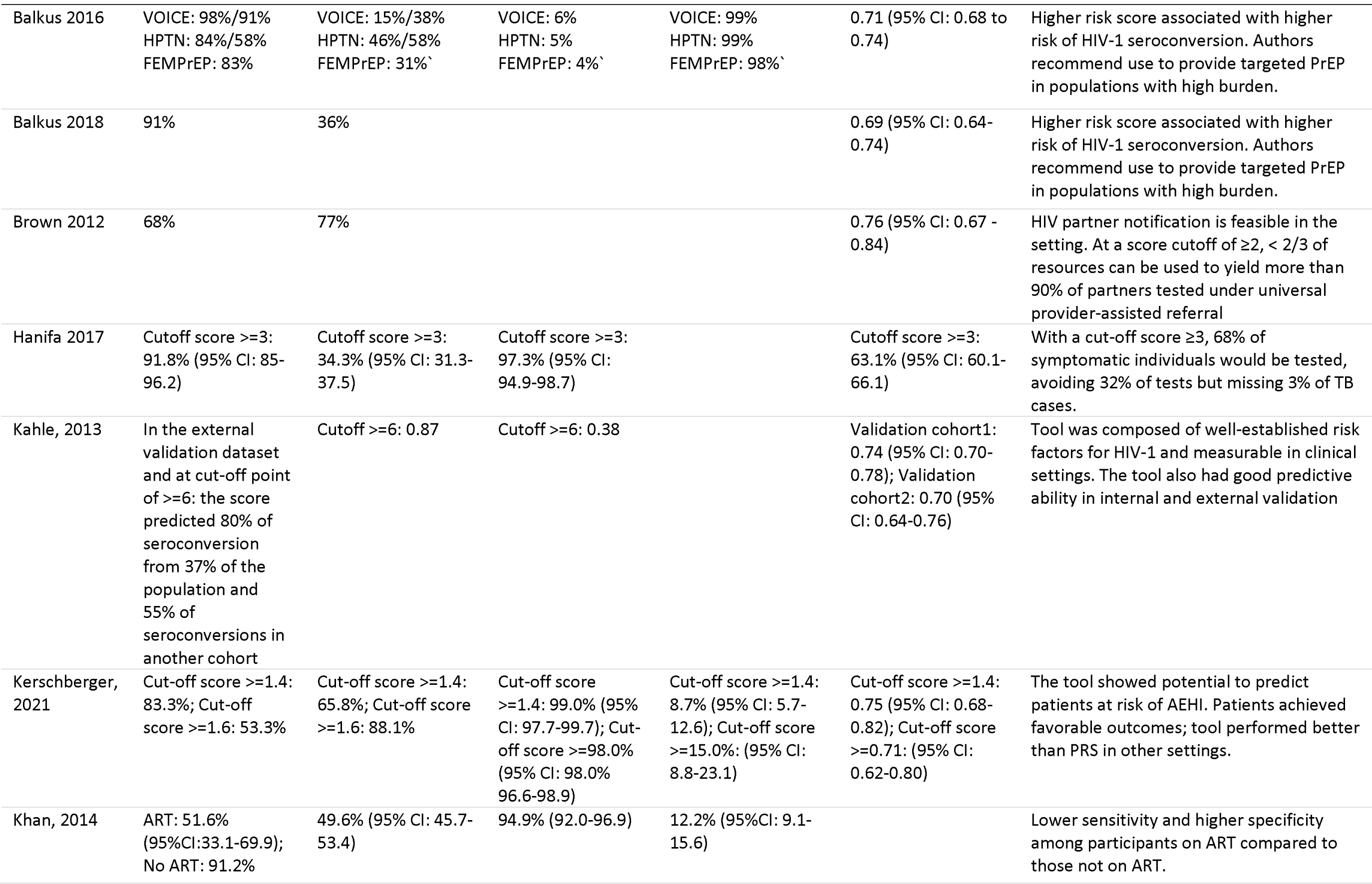
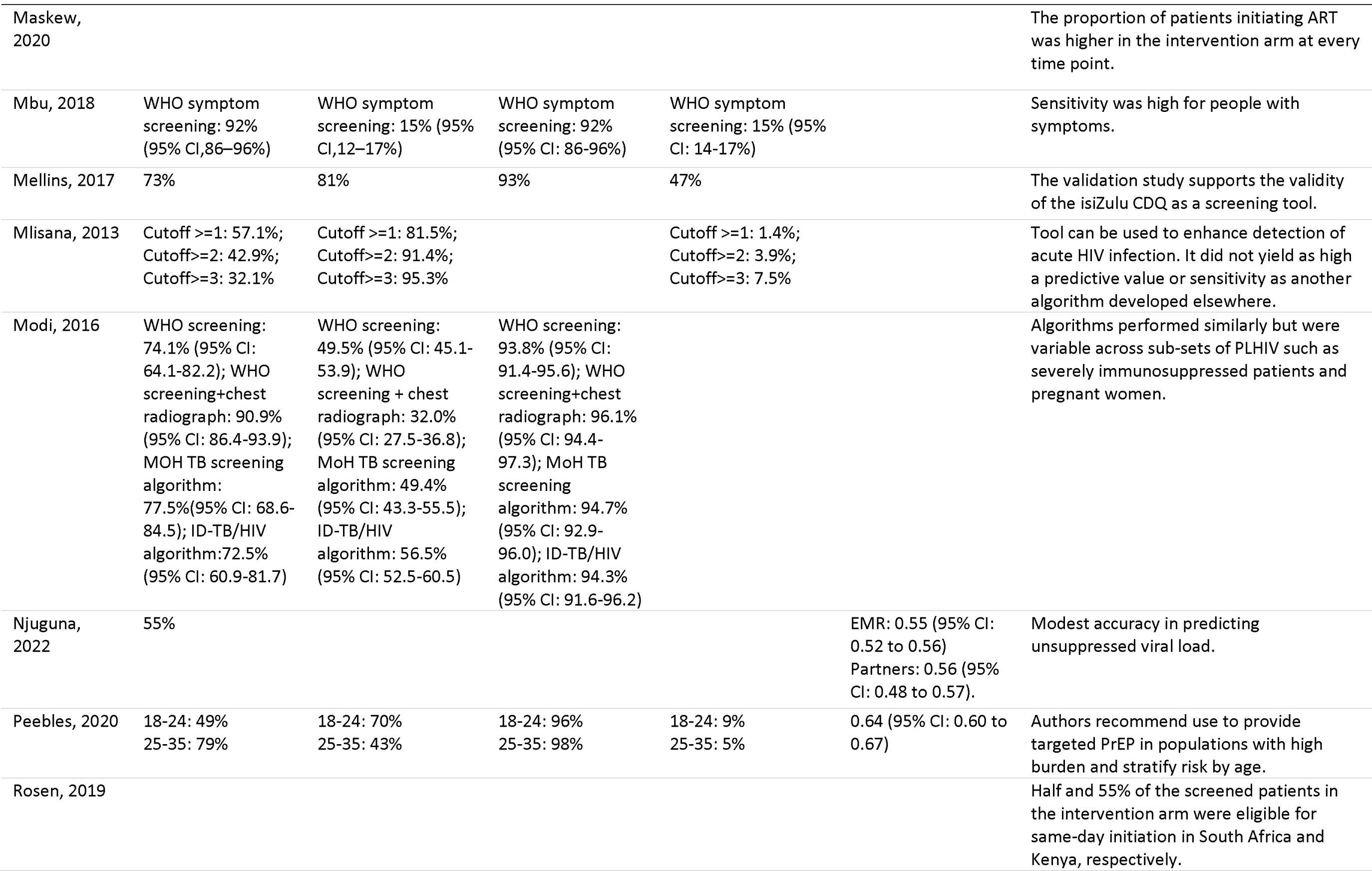
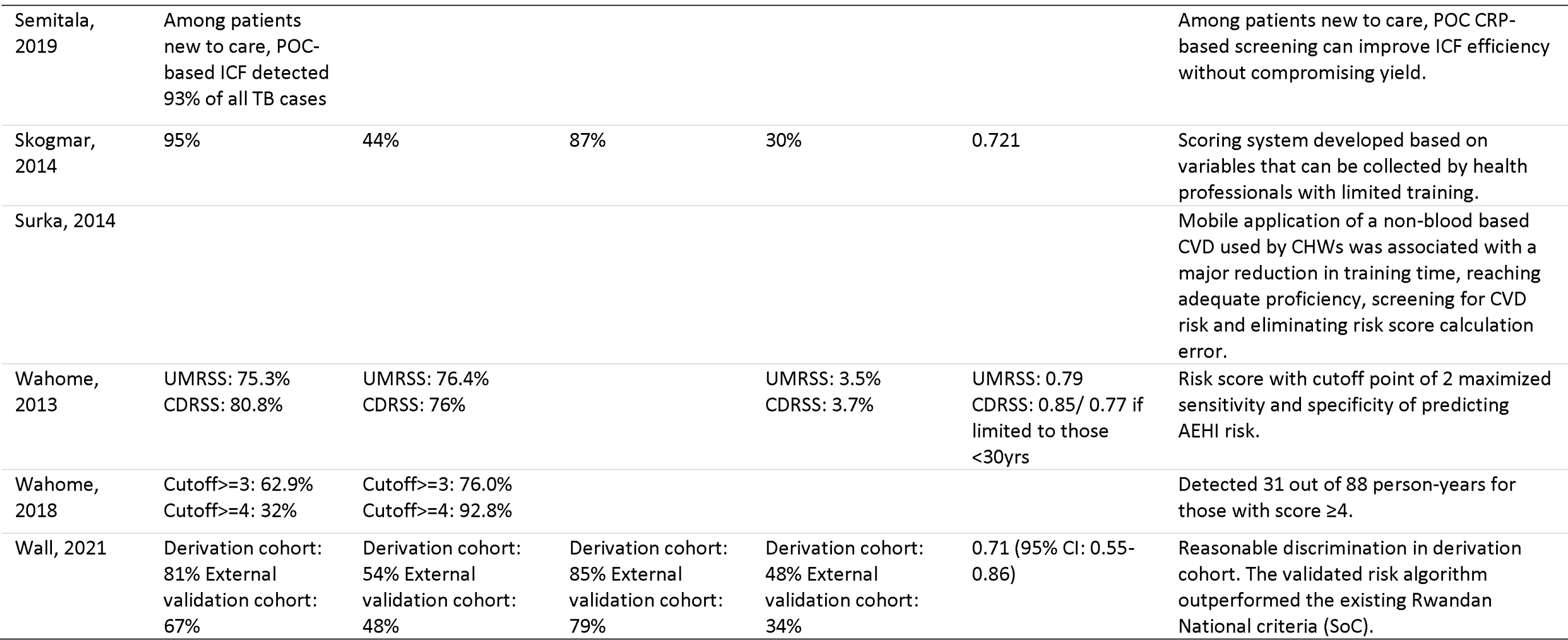
Performance metrics of risk scoring tools.

Figure 3 illustrates the summary performance of tools evaluated in the included studies. For each tool, we plotted a solid line connecting the point estimate for sensitivity with the estimate for false positive rate (1-specificity). Next NPV, PPV, and AUC (where available) were plotted as individual points around the plotted line. In this way, a tool with high performance metrics would be visualised as a wide solid line connecting point estimates for sensitivity and false positive rate with AUC and other metrics clustered near the right side of the axis. Several tools demonstrated these patterns[32,36]. There was a frequently observed trade-off, however, between sensitivity and specificity. Three TB screening algorithms assessed in Kenya, for example, demonstrated sensitivity between 73-91%, while specificity was <50% for all three [50]. The effect of outcome prevalence on PPV and NPV was also observed, such as for the VOICE risk scoring tool predicting risk of HIV acquisition[39].

**Figure 3.**
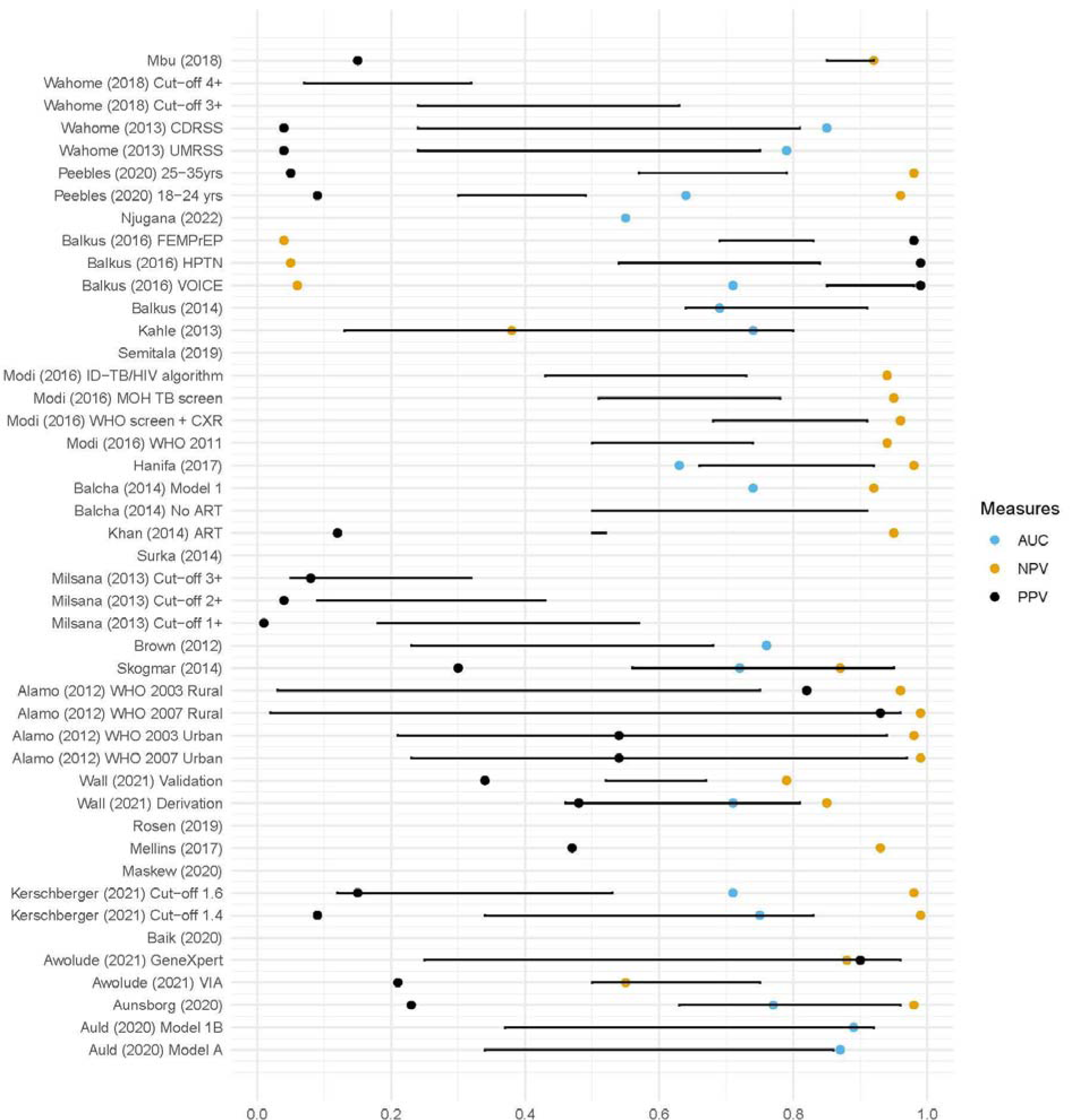
Summary performance metrics of risk triaging tools*. *For each study, the solid line represents a plotted line connecting the point estimate for sensitivity with the point estimate for false positive rate (1-specificity) for that specific tool. Where available, negative predictive value (NPV), positive predictive value (PPV), and area under the curve (AUC) were plotted as individual points around the plotted line as indicated in the legend.

### Health system effects

As noted previously, 14 of the 28 studies included in this review did not implement the risk triaging tools they developed. The remaining 14 implemented the tools in patient cohorts, either in clinical trial (n=4) or observational (n=10) settings. Few of the studies reported effects of tool implementation beyond the accuracy of the risk predictions made. We found no discussion of health system effects as defined in our outcomes in 4 of the implementation studies. Other studies posited (but did not provide evidence of) potential impacts, should their tool be implemented at scale. Comments on implementation fell into 3 categories: 1) the potential for safely shifting screening services to lower cadre staff, even if higher cadre staff are needed to provide treatment; 2) improving efficiency and reducing service delivery costs; and 3) the use of existing or easily collected indicators, without the need for laboratory tests or equipment that is not typically available in primary clinics. None was able to provide tangible evidence of potential impact of the tool in question.

Studies that reported potential improved efficiency and cost savings with risk triaging attributed such gains to fewer patients requiring further investigation after risk screening[33]; task shifting as less expensive cadres including lay health workers conduct the screening[42,48,56]; targeting post-risk screening care to patient groups with the highest benefit[38,54]; and reduced resources required for patient tracing due to improved yield[41]. Studies that both implemented and developed risk scoring tools further reported potential health system benefits through portability, ease of transferability[37], and scalability of risk scoring tools in resource limited settings when compared to advanced microbiological diagnostics[35]. They also noted improved staff alertness of the condition in question when a risk scoring tool is actively integrated in routine clinic processes[35]. In some cases, other indirect benefits to implementation of triaging tools were observed, such as general improvements in awareness and uptake of TB screening reported after implementation of the TBscore tool[35].

### Assessment of risk of bias

We evaluated each included article for risk of bias utilising the JBI checklist for diagnostic test accuracy which considers important potential areas of bias for the measurement and interpretation of test accuracy results [31]. This includes features of study design that impact test metrics such as avoiding a case-control approach (in which the people with the disease (cases) are selected from a different population than the control persons without the disease) in favour of cross-sectional or cohort designs in which all participants undergo the test or triaging process. The latter approach reflects reality better than the case–control design and is more likely to provide valid estimates of diagnostic accuracy [60]. Overall, the median bias score was 12% (IQR 0-25%). Nineteen studies were classified as low risk of bias and 9 with moderate risk of bias. No articles indicated high risk of bias. Details are shown in S4 Table.

## DISCUSSION

In settings that face a scarcity of resources—a situation that prevails in many public sector healthcare settings globally—identifying individuals at higher risk of negative outcomes now or in the future is a critical step in ensuring that available resources are allocated where they can do most good. While risk triaging is undoubtedly conducted informally by most if not all healthcare providers, based on their own experience and judgment, widespread use of clinical triaging tools appears to be rare in primary healthcare clinic settings in sub-Saharan Africa. There is even less use of structured instruments to identify future behavioural risks, such as attrition from care. In resource-scarce settings, many facilities are high-volume, staff-constrained sites, where effective triaging tools could improve the quality of clinical care while also increasing the efficiency of health care resource utilization.

The systematic review reported here is, to our knowledge, the first attempt to synthesize experience in developing, testing, and, in a few cases, implementing risk triaging tools in these settings. Our findings offer a mixed message, in terms of the future role of risk triaging in routine care. On one hand, most of the tools we identified were at least somewhat successful in identifying potential risk, with the majority of tools correctly identifying the outcome of interest in upwards of 70% of screened patients. The relatively low specificity of most of the tools is not a serious concern, since most were developed as screening tools rather than diagnostics, making high sensitivity the priority. For purposes of identifying groups of patients who are likely to benefit from additional testing or monitoring, versus those who clearly have no need for additional services, even a tool with modest sensitivity and low specificity has the potential to improve resource allocation, particularly in settings where prevalence of the outcome or condition screened for is high. In addition, an encouraging proportion of the tools we reviewed were explicitly designed to rely on indicators readily available at the primary healthcare level, rather than laboratory tests or imaging assays, though many also emphasized risk factors that are not easily addressed through practical interventions, such as age and sex.

On the other hand, based on the published reports, very few of the risk triaging tools identified were ever implemented beyond their initial studies or scaled up outside the research setting. Some developed and internally validated their risk scores using retrospective data and some tested risk triaging tools in small study populations, but none reported routine adoption of the prediction tools. There are likely multiple reasons for this, ranging from lack of logistical feasibility (e.g. an instrument required too much data to be feasible to administer during a routine clinical encounter) to lack of dissemination on the part of the developers. Without a clearly demonstrated need and demand for a triaging approach, as well as motivated and empowered champions willing and able to advocate for scale-up, many potentially well-performing and useful tools will not be utilised beyond their development setting. The lack of published information about how improvements to routine procedures, such as a new risk triaging tool, can be adopted at large scale is a gap in our understanding of how to improve primary healthcare performance.

While few of the tools we reviewed appear to have been adopted for widespread use, there is one (and perhaps only one) that has: the WHO’s TB symptom screening tool. The four- or five-item screening questionnaire is in widespread, if not near-universal, use throughout sub-Saharan Africa for people living with HIV. Several of the studies included here evaluated the TB symptom screen, alone or in combination with other indicators, and generated mixed results in terms of sensitivity and specificity for different populations. Exploration of how the TB symptom screen came to be so widely accepted and utilized suggests that a combination of meta-analysis of data from many sources, early and substantial engagement of policy makers, and international agency leadership allowed the TB symptom screen first to be incorporated into WHO guidelines[61] and then integrated into national guidelines and practice throughout the region. Better understanding of this process may be useful in introducing other risk triaging tools into widespread use.

Half of the studies included in our review addressed risks associated with HIV, whether transmission, treatment initiation, viral suppression, or mortality, and another third focused on TB case-finding. This may be explained by the very large numbers of individuals requiring these services in many sub-Saharan African countries. It may also reflect the large amounts of funding that have been available for HIV- and TB-related research and interventions in general, compared to other conditions that could be amenable to risk triaging, such as NCDs. Interestingly, however, none of the HIV-related studies included here offered a risk triaging tool aimed directly at adherence to HIV treatment, with retention in care as the outcome of interest. Researchers may have had more success in assessing risks of HIV acquisition, readiness for treatment, viral suppression, and mortality than in finding consistent risk predictors of the broader outcome of “retention,” but it is retention that remains the single most difficult challenge for national HIV programs. Continued research in this area may thus be warranted.

The studies reviewed here do provide a number of general and specific lessons about the characteristics of a promising risk triaging tool and how such a tool may be adopted into widespread use. First, tools must be brief and easy to integrate into routine care without disrupting established practices and rely on risk factors that are either already routinely recorded or are easy and quick to collect. Expanding existing data collection requirements is simply not feasible in busy, understaffed clinics where computer access, and even electricity, cannot be taken for granted. In the authors’ experience, moreover, primary healthcare clinic staff may resist innovations that they believe will increase their workload or complicate their tasks, even if the ultimate effect may be the opposite, such as an intervention that increases the number of candidates for early screening but reduces long-term treatment demand. Convincing healthcare staff that a new tool is worth the effort is likely to be much more successful if the tool itself is simple to understand and implement. Tools that can be implemented, wholly or in part, by lay cadres such as counsellors, peer supporters or community healthcare workers, rather than solely by scarcer cadres such as nurses, may also be more promising for widespread adoption, particularly as provision of primary care move towards community-based and other out-of-facility service delivery models.

Second, where risk scores are generated and utilised, interpretation of the output scores should be simple and straightforward. Decision cut-off points (e.g. having a risk score above or below a threshold) must be clear and well justified. We also note that while none of the studies we reviewed discussed offering healthcare providers discretion in interpreting risk scores, it seems reasonable that these scores should be seen as guidelines rather than absolute rules. If a clinician interacting with a patient believes that the risk score generated does not lead to the correct management pathway, clinician judgment may be preferred, as it is in many clinical situations. Similarly, individual patients may have strong reasons for preferring one pathway to another. Since patient cooperation is essential to achieving good outcomes, such preferences should be taken into account, no matter what a risk triaging tool suggests.

Third, in most cases, assessing risk is not an end in itself. It does little good, for example, to designate an HIV treatment patient as having a low or high risk of treatment interruption if there is not a clear pathway for responding to the conditions that create the assessed level of risk. A client who is determined to be at high risk of disengagement due to fears of disclosure and stigma will require an entirely different intervention strategy from a client at high risk of disengagement due to an employment situation that does not allow clinic visits during working hours. Risk triaging tools should thus come with intervention strategies or programs designed for the specific risk factors that contribute to the score or other outcome.

Fourth, performance metrics should not be the sole decision-making criterion determining implementation of a potential tool. A tool with low specificity may be acceptable if the purpose of risk triaging is to reduce the population of patients who may require additional diagnosis, care, or support, because even a relatively non-specific tool has the potential to identify those patients who do <u>not</u> require additional services. In any group of 100 clients initiating ART, for example, it is likely that at least 60, if not more, are at very low risk of disengagement from care in their first year [14,62]. Standard of care guidelines in many settings would require all one hundred of them to be assigned multiple adherence counselling sessions in the months after initiation. A triaging tool that could identify even half (30) of the true “low risk” patients would allow the facility to focus counselling resources more efficiently among those who are most likely to derive benefit from such an intervention. While not ideal, this would still represent a potentially valuable improvement in facility performance.

Finally, no risk triaging tool should be expected to perform consistently across all populations and settings. Setting matters when evaluating tool performance, even for a screening tool for TB risk, which relies on presumably objective symptom and laboratory indicators[63] and will be affected by TB prevalence rates in the setting the tool is implemented. For a behavioral risk tool, community and facility factors may be as important in predicting risk as individual characteristics. Travelling long distances to an ART clinic, for example, is frequently identified as a barrier to treatment adherence, and many programs offer community-based service delivery to address this barrier. In communities where stigma around HIV is high, however, individuals requiring HIV care may in fact prefer to travel further to facilities based in communities where they are less likely to be recognized by those to whom they have not disclosed their status. Local refinement of established risk triaging instruments may thus be desirable.

This review had a number of limitations. First, our search terms excluded all inpatient settings. This may have systematically excluded entire medical conditions that frequently use risk triaging approaches, such as disease due to the Ebola virus [64]. Second, while we included terms for performance test metrics such as sensitivity, specificity, and accuracy to increase the yield of studies reporting methodological results, we acknowledge that the terminology around test metrics is not universally standard and may have led to relevant studies being missed, particularly among older studies. Third, though we were unable to address this limitation through search methodology, we acknowledge that a systematic review will not observe tools that have not been described and published in academic literature. There may be some inherent reporting bias if results of tools that did not work were less likely to be published, but it is also possible that implementing partners and organizations have designed and implemented successful triaging tools and systems in primary health contexts but not evaluated or published these. Fourth, as discussed above, generalizability of the performance of any given risk triaging tool is uncertain. Tools that rely on clinical symptoms or test results may be applicable across multiple settings and populations, but tools that assess behavioural risks seem likely to be highly context-specific. Finally, in determining which papers to include in the analysis, the distinction between risk triaging and diagnosis was sometimes unclear and required author discretion; it is possible that some sources that others may regard as examples of risk triaging were categorized as diagnosis and excluded.

## Conclusion

Despite the limitations listed above, our review highlights some important considerations for the implementation of risk triaging tools in the context of HIV retention. Many studies have achieved relatively impressive performance in identifying the risks they target within the data sets or small patient cohorts in which they were designed and validated. The lack of large-scale adoption and implementation of any of these instruments, however, points to serious gaps in previous efforts. Bringing implementers, policy makers, funders, and potentially clients themselves into the process of identifying needs, developing tools and intervention strategies, and assessing feasibility may be an essential component of improving health outcomes through risk stratification.

## Data Availability

All data produced in the present work are contained in the manuscript.

## Ethics approval and consent to participate

This is a systematic review of published information and no direct contact with participants or access to participant data beyond what is reported in the published studies occurred.

## Availability of data and materials

All data used in this paper are included in the cited manuscripts and publicly available in the cited sources.

## Competing interests

The authors declare that they have no competing interests.

## Funding

Funding for the study was provided the Bill & Melinda Gates Foundation through INV-031690 to Boston University. The funder had no role in study design, data collection and analysis, decision to publish, or preparation of the manuscript.

## Author contributions

MM, SR, and LS conceived of and designed the study. DF defined and ran the search. MM, LS, and MB identified and reviewed sources and extracted data. MM, LS, and MB analyzed the data and drafted the manuscript. All authors reviewed and edited the manuscript.

## Acknowledgements

None

## Supporting information

S1 Text. Review protocol S1 Table. Search strategy

S2 Table. Search terms-PubMed S3 Table. Prisma checklist.

S4 Table. Summary of bias using the Joanna Briggs Checklist for Diagnostic Test Accuracy

## Notes

### Competing Interest Statement

The authors have declared no competing interest.

### Clinical Protocols

https://www.crd.york.ac.uk/prospero/display_record.php?RecordID=328209

