## Supplemental file 1 for "Implementation of risk triaging in primary healthcare facilities in Sub-Saharan Africa: A systematic review"

**Project question**

Review whether and how risk-triaging has previously been used in primary healthcare clinics in Africa and the features that make this approach most likely to succeed. Summarize findings about the potential value of risk triaging in a project report and, if warranted, a journal publication.

**Introduction**

**Rationale**

UNAIDS set fast-track targets aimed at ending AIDS as a public health threat by 2030. The targets are to ensure that by 2030, 95% of people living with HIV (PLHIV) are aware of their status, 95% of PLHIV who know their status are on treatment and 95% of PLHIV on treatment have suppressed viral load (1). Suppressed viral load among PLHIV is important to ensuring strong immunity and reduced HIV transmission (1). First six months of antiretroviral therapy (ART) initiation are especially important to patient outcomes including viral load suppression (2,3).

Expansion of antiretroviral therapy (ART) in sub-Saharan Africa (SSA) and the recommendation of placing all PLHIV on treatment regardless of clinical status, has improved patient outcomes and moved countries closer to the UNAIDS fast-track targets (2,4). On the flip side, the policy has also contributed to patient loss to follow-up (5) by transferring patients loss from before to after treatment initiation.

HIV programmes have been facing challenges retaining patients on ART (2,6–12). Reported rates of ART retention in the first 12 months of initiation vary widely across settings (3,9,13–15). Studies in Malawi and Zimbabwe reported retention rates of 80.7% and 86.5% in the first 12 months, respectively (6,16). A study in Kenya reported a third of patients not retained into care in the first 12 months (9). The World Health Organization (WHO) summarized retention rates as ranging from 64% to 94% (15).

A combination of social, economic and structural factors are responsible for challenges in retaining patients in ART programmes (10). Such factors include; age, sex, advanced disease, stigma, high user costs including mobility challenges to facilities, treatment fatigue and long waiting times (3,5,6,9,14,17–19).

Identifying early patients likely to be lost to follow -up can help improve patient retention in care by allowing for differentiated care and other adherence interventions (5,20,21). Prediction or risk triaging tools can be used to identify such patients. Such tools have been developed or used previously to identify: patients at the risk of ART default and poor viral load outcomes (5,20–22); patients qualifying for same day ART initiation (23,24); adults and children in need of HIV testing (25,26); patients likely to return to care after disengagement (27); to prioritize patients in need of TB screening for timely treatment (28–30); to identify the risk of non-adherence to treatment (31); and, to reduce patient waiting times in clinics (32).

We seek to conduct a systematic review of the use of such risk triaging tools in SSA. The findings will be used to develop a risk triaging tool for ART default with the aim of providing differentiated care for patients with a high risk of default in the first 6 months of initiation. We will answer the following additional questions:

i. How has risk triaging been applied in primary healthcare facilities in SSA?
ii. What features make risk triaging more likely to succeed?

**Methods**

**Population**: Adults aged 18 or above seeking care in primary healthcare facilities in SSA
**Intervention**: Implementation of risk triaging in primary healthcare facilities in SSA
**Comparator**: No comparator as this will be a descriptive review
**Primary outcomes:**

1. Patient outcomes as a result of the implementation of a risk triaging tool
   i. Effect on health outcomes
   ii. Effect on efficiency
      - Patient healthcare facility experience such as, waiting times
      - Provider performance through e.g. task shifting
2. Risk triaging tool performance
   i. Sensitivity
   ii. Specificity
   iii. Negative predictive values
   iv. Positive predictive value

**Secondary outcomes:**

1. Risk triaging tool implementation process
   i. How was the tool implemented?
      o Electronic or paper-based
      o Checklist/colour codes/series of questions
   ii. Which section of the clinic implemented the tool?
   iii. Who delivered the tool?
      o Staff cadre
   iv. At what point in the patient flow was the risk triaging tool applied?
2. Risk triaging tool contents
   i. How many items were included in the tool?
   ii. How were the items derived?
   iii. How was the tool validated?
3. Costs and cost-effectiveness of risk triaging tools in SSA
   i. costs and cost-effectiveness of the risk triaging tool
4. Experience scaling up such tools
   i. Sustainability of a scaled-up risk triaging tool

**Study eligibility criteria**

We will include reports with primary, patient-level data from retrospective or prospective cohorts collected under any study design (trial, observational) with or without a comparison group, systematic reviews, meta-analyses. Case series or reports, purely qualitative studies, treatment guidelines, mathematical models, editorials, commentaries, study or trial protocols will be excluded.

All included studies will be reporting and/or evaluating a risk triaging tool or algorithm used for adults aged 18 or above in primary care facilities in SSA. White papers and published protocols with no data will be excluded.

**Information sources**

We will search for published studies and grey literature in MEDLINE, Cochrane Library, Web of Science, Embase, Scopus, and ClinicalTrials.gov.

**Search strategy**

Table 1 presents the proposed search strategy. Results will be limited to English language and only studies published from the year 2018 will be included.

**Table 1: Proposed search strategy**

| | |
|---|---|
| Population | (adult[MeSH] OR " adult population"[MeSH]<br><br>**AND**<br>("Primary health care" OR "PHC" OR "Community Health" OR "Outpatient care" OR "Outpatient" OR "Public health" OR "Public-sector primary care" OR "primary care facility" OR "Care, Primary Health" OR "Health Care, Primary" OR "Primary Healthcare" OR "Healthcare, Primary" OR "Primary Care" OR "Care, Primary" OR "Community Health Service" OR "Health Service, Community" OR "Service*, Community Health" OR "Health Services, Community" OR "Community Health Care" OR "Care, Community Health" OR "Health Care, Community" OR "Community Healthcare*" OR "Healthcare*, Community" OR "Care, Ambulatory"OR "Care, Outpatient"OR "Health Service*, Outpatient" OR "Outpatient Health Service*" OR "Service, Outpatient Health" OR "Outpatient Service*" OR "Services, Outpatient Health" OR "Urgent Care" OR "Care*, Urgent" OR "Clinic Visit*" OR "Visit*, Clinic)) |
| Intervention | ("Risk triaging" OR "Risk triag*" OR "Risk scor*" OR "Algorithm" OR "Predictive algorithm" OR "Predictive score" OR "Predictive model" OR "Risk model" OR "Patient screen" OR "Risk screening" OR "Score, Risk" OR "Risk Factor Score*" OR "Score, Risk Factor*"<br><br>**AND**<br>"Factor, Time" OR "Time Factor*" OR "Time Series" OR Longitudinal OR "Longitudinal Stud*" OR "Stud*, Longitudinal" OR "Follow Up Stud*" OR Follow-Up Stud*" OR "Stud*, Follow-Up" OR "Followup Stud*" OR "Stud*, Followup") |
| Outcomes | ("Clinical outcome" OR "Treatment success" OR "Retention in care" OR "Re-engagement in care" OR Mortality OR "Clinic performance" OR "Clinic waiting times" OR "waiting times"OR "Resource utilization" OR "cost" OR "Resource utilisation" OR "Health Resource" OR "Resource, Health" OR "Resources, Health" OR Resources OR Resource OR "Program outcome" OR "Task shifting")<br><br>**OR**<br>("Performance metrics" OR "Test metrics" OR Sensitivity OR Specificity OR "Positive predictive value" OR "Negative predictive value" OR "Area under the curve" OR "Random operator curve" OR "Accuracy" OR "Precision") |

| | |
|---|---|
| Context | (Africa[MeSH:noExp] OR Sub-Saharan-Africa* OR Subsaharan-Africa* OR Africa South of the Sahara[MeSH] OR Central-africa* OR Eastern-africa* OR East-africa* OR Southern-africa* OR South-africa* OR Western-africa* OR West-africa* OR Cameroon* OR Central-african-republic* OR Chad* OR Congo* OR DRC OR Equatorial-guinea* OR Gabon* OR Sao-Tome-and-Principe* OR Burundi* OR Djibouti* OR Eritrea* OR Ethiopia* OR Kenya* OR Rwanda* OR Somalia* OR South-sudan* OR Sudan* OR Tanzania* OR Uganda* OR Angola* OR Botswana* OR Eswatini* OR Swaziland* OR Lesotho* OR Malawi* OR Mozambique* OR Namibia* OR Zambia* OR Zimbabwe* OR Benin OR Burkina-Faso* OR Cabo-Verde* OR Cote-d'Ivoire* OR Ivory-Coast* OR Gambia* OR Ghana* OR Guinea* OR Guinea-Bissau* OR Liberia* OR Mali* OR Mauritania* OR Niger* OR Nigeria* OR Senegal* OR Sierra-Leone* OR Togo*) |

**Data management**
Study records will be imported into an Endnote reference library. Titles and abstracts will be screened using Rayyan and thereafter exported to a Mendeley reference library for full text screening. Three independent reviewers will be involved in all process of the review. LS will first screen the titles and abstracts with VN and MM serving as second reviewers. Disagreements will be resolved through discussion among the three reviewers.

**Data items to be extracted from studies**
We will extract descriptions of patients, timing of the risk triaging intervention in relation to provision of primary healthcare, facility location and type, service delivery models and services provided. We will further extract descriptions of the risk triaging tool and its performance, impact on patient outcomes and providers' performance, data on the tool's scalability, costs, and cost-effectiveness. and application of the tool at scale.

**Risk of bias**
Bias will be assessed using the Joanna Briggs Institute (JBI) critical appraisal checklist for systematic reviews. The JBI checklist asks a set of 11 closed ended questions for every study included in a review to evaluate any bias in the design, conduct and analysis in the studies meeting the inclusion criteria (33).

**Data synthesis**
This review will be descriptive and therefore, there will be no meta-synthesis of data conducted. We will however where feasible, group the data by themes such as disease and similarity in contents or implementation of risk triaging tools.

**Confidence in cumulative evidence**
Summary of papers included in addition to the quality of individual papers will be presented using the Grading of Recommendations, Assessment, Development and Evaluations (GRADE) tool (34). Following the GRADE tool, studies will be rated based on risk of bias, imprecision, inconsistency, indirectness and publication bias.
