## Supplemental tables for "Implementation of risk triaging in primary healthcare facilities in Sub-Saharan Africa: A systematic review"

**S1 Table. Search strategy**

| **Parameter** | **Inclusion criteria** | **Exclusion criteria** |
| --- | --- | --- |
| Population | Ages 18+ years; accessing primary health care in a public-sector setting | Paediatric and adolescent populations aged <18 years; accessing care in the private sector |
| Geographic region | Sub-Saharan Africa | None |
| Intervention | Implementation of risk triaging in primary healthcare settings | None |
| Study design | Reports primary, patient-level data from retrospective or prospective cohorts collected under any study design (trial, observational) with or without a comparison group; systematic reviews, meta-analyses | Case series or reports, purely qualitative studies, treatment guidelines, mathematical models, editorials, commentaries, study or trial protocols |
| Required descriptive data | Describes all of patients, location, timing of risk triaging intervention in relation to provision of primary health care, facility type, service delivery models and services provided to the public sector through government-managed public health infrastructure or through NGO/private programs or facilities that serve the uninsured sector | Insufficient description of all the characteristics needed to describe the study population and outcome |
| Comparator | None required – descriptive review | None |
| Outcomes (primary) | 1.Patient outcomes as a result of the implementation of a risk triaging tool  2.Risk triaging tool performance metrics | Insufficient detail provided to estimate outcome |
| Timing | A majority of data collected after January 1, 2017; censored at search date of July 25, 2022 | A majority of data accrued before January 1, 2017; published after censoring date of July 25, 2022 |

**S2 Table. Search terms – PubMed**

| Population | (adult[MeSH] OR " adult population"[MeSH]  **AND**  (“Primary health care” OR “PHC” OR “Community Health” OR “Outpatient care” OR “Outpatient” OR “Public health” OR “Public-sector primary care” OR “primary care facility” OR “Care, Primary Health” OR “Health Care, Primary” OR “Primary Healthcare” OR “Healthcare, Primary” OR “Primary Care” OR “Care, Primary” OR “Community Health Service” OR “Health Service, Community” OR “Service*, Community Health” OR “Health Services, Community” OR “Community Health Care” OR “Care, Community Health” OR “Health Care, Community” OR “Community Healthcare*” OR “Healthcare*, Community” OR “Care, Ambulatory”OR “Care, Outpatient”OR “Health Service*, Outpatient” OR “Outpatient Health Service*” OR “Service, Outpatient Health” OR “Outpatient Service*” OR “Services, Outpatient Health” OR “Urgent Care” OR “Care*, Urgent” OR “Clinic Visit*” OR “Visit*, Clinic)) |
| --- | --- |
| Intervention | (“Risk triaging” OR “Risk triag*” OR “Risk scor*” OR “Algorithm” OR “Predictive algorithm” OR “Predictive score” OR “Predictive model” OR “Risk model” OR “Patient screen” OR “Risk screening” OR “Score, Risk” OR “Risk Factor Score*” OR “Score, Risk Factor*”  **AND**  “Factor, Time” OR “Time Factor*” OR “Time Series” OR Longitudinal OR “Longitudinal Stud*” OR “Stud*, Longitudinal” OR “Follow Up Stud*” OR Follow-Up Stud*” OR “Stud*, Follow-Up” OR “Followup Stud*” OR “Stud*, Followup”) |
| Outcomes | (“Clinical outcome” OR “Treatment success” OR “Retention in care” OR “Re-engagement in care” OR Mortality OR “Clinic performance” OR “Clinic waiting times” OR “waiting times”OR “Resource utilization” OR “cost” OR “Resource utilisation” OR “Health Resource” OR “Resource, Health” OR “Resources, Health” OR Resources OR Resource OR “Program outcome” OR “Task shifting”)  **OR**  (“Performance metrics” OR “Test metrics” OR Sensitivity OR Specificity OR “Positive predictive value” OR “Negative predictive value” OR “Area under the curve” OR “Random operator curve” OR “Accuracy” OR “Precision”) |
| Context | (Africa[MeSH:noExp] OR Sub-Saharan-Africa* OR Subsaharan-Africa* OR Africa South of the Sahara[MeSH] OR Central-africa* OR Eastern-africa* OR East-africa* OR Southern-africa* OR South-africa* OR Western-africa* OR West-africa* OR Cameroon* OR Central-african-republic* OR Chad* OR Congo* OR DRC OR Equatorial-guinea* OR Gabon* OR Sao-Tome-and-Principe* OR Burundi* OR Djibouti* OR Eritrea* OR Ethiopia* OR Kenya* OR Rwanda* OR Somalia* OR South-sudan* OR Sudan* OR Tanzania* OR Uganda* OR Angola* OR Botswana* OR Eswatini* OR Swaziland* OR Lesotho* OR Malawi* OR Mozambique* OR Namibia* OR Zambia* OR Zimbabwe* OR Benin OR Burkina-Faso* OR Cabo-Verde* OR Cote-d'Ivoire* OR Ivory-Coast* OR Gambia* OR Ghana* OR Guinea* OR Guinea-Bissau* OR Liberia* OR Mali* OR Mauritania* OR Niger* OR Nigeria* OR Senegal* OR Sierra-Leone* OR Togo*) |

**S3 Table. Prisma checklist**

| **Section and Topic** | **Item #** | **Checklist item** | **Location where item is reported** |
| --- | --- | --- | --- |
| **TITLE** | | |  |
| Title | 1 | Identify the report as a systematic review. | Title page |
| **ABSTRACT** | | |  |
| Abstract | 2 | See the PRISMA 2020 for Abstracts checklist. | Page 2 |
| **INTRODUCTION** | | |  |
| Rationale | 3 | Describe the rationale for the review in the context of existing knowledge. | Page 3 |
| Objectives | 4 | Provide an explicit statement of the objective(s) or question(s) the review addresses. | Page 4 |
| **METHODS** | | |  |
| Eligibility criteria | 5 | Specify the inclusion and exclusion criteria for the review and how studies were grouped for the syntheses. | Pages 5-6, S1 Table |
| Information sources | 6 | Specify all databases, registers, websites, organisations, reference lists and other sources searched or consulted to identify studies. Specify the date when each source was last searched or consulted. | Page 6 |
| Search strategy | 7 | Present the full search strategies for all databases, registers and websites, including any filters and limits used. | S2 Table |
| Selection process | 8 | Specify the methods used to decide whether a study met the inclusion criteria of the review, including how many reviewers screened each record and each report retrieved, whether they worked independently, and if applicable, details of automation tools used in the process. | Page 6 |
| Data collection process | 9 | Specify the methods used to collect data from reports, including how many reviewers collected data from each report, whether they worked independently, any processes for obtaining or confirming data from study investigators, and if applicable, details of automation tools used in the process. | Page 6 |
| Data items | 10a | List and define all outcomes for which data were sought. Specify whether all results that were compatible with each outcome domain in each study were sought (e.g. for all measures, time points, analyses), and if not, the methods used to decide which results to collect. | Page 7 |
|  | 10b | List and define all other variables for which data were sought (e.g. participant and intervention characteristics, funding sources). Describe any assumptions made about any missing or unclear information. | Page 7 |
| Study risk of bias assessment | 11 | Specify the methods used to assess risk of bias in the included studies, including details of the tool(s) used, how many reviewers assessed each study and whether they worked independently, and if applicable, details of automation tools used in the process. | Page 7 |
| Effect measures | 12 | Specify for each outcome the effect measure(s) (e.g. risk ratio, mean difference) used in the synthesis or presentation of results. | Pages 7-8 |
| Synthesis methods | 13a | Describe the processes used to decide which studies were eligible for each synthesis (e.g. tabulating the study intervention characteristics and comparing against the planned groups for each synthesis (item #5)). | n.a.(page 8) |
|  | 13b | Describe any methods required to prepare the data for presentation or synthesis, such as handling of missing summary statistics, or data conversions. | n.a. |
|  | 13c | Describe any methods used to tabulate or visually display results of individual studies and syntheses. | n.a. |
|  | 13d | Describe any methods used to synthesize results and provide a rationale for the choice(s). If meta-analysis was performed, describe the model(s), method(s) to identify the presence and extent of statistical heterogeneity, and software package(s) used. | n.a. |
|  | 13e | Describe any methods used to explore possible causes of heterogeneity among study results (e.g. subgroup analysis, meta-regression). | n.a. |
|  | 13f | Describe any sensitivity analyses conducted to assess robustness of the synthesized results. | n.a. |
| Reporting bias assessment | 14 | Describe any methods used to assess risk of bias due to missing results in a synthesis (arising from reporting biases). | Page 8 |
| Certainty assessment | 15 | Describe any methods used to assess certainty (or confidence) in the body of evidence for an outcome. | n.a. |
| **RESULTS** | | |  |
| Study selection | 16a | Describe the results of the search and selection process, from the number of records identified in the search to the number of studies included in the review, ideally using a flow diagram. | Pages 8-9, figure 1 |
|  | 16b | Cite studies that might appear to meet the inclusion criteria, but which were excluded, and explain why they were excluded. | Page 9 |
| Study characteristics | 17 | Cite each included study and present its characteristics. | Table 1, Figure 2 |
| Risk of bias in studies | 18 | Present assessments of risk of bias for each included study. | S4 Table |
| Results of individual studies | 19 | For all outcomes, present, for each study: (a) summary statistics for each group (where appropriate) and (b) an effect estimate and its precision (e.g. confidence/credible interval), ideally using structured tables or plots. | Table 2, Figure 3 |
| Results of syntheses | 20a | For each synthesis, briefly summarise the characteristics and risk of bias among contributing studies. | n.a. |
|  | 20b | Present results of all statistical syntheses conducted. If meta-analysis was done, present for each the summary estimate and its precision (e.g. confidence/credible interval) and measures of statistical heterogeneity. If comparing groups, describe the direction of the effect. |  |
|  | 20c | Present results of all investigations of possible causes of heterogeneity among study results. |  |
|  | 20d | Present results of all sensitivity analyses conducted to assess the robustness of the synthesized results. |  |
| Reporting biases | 21 | Present assessments of risk of bias due to missing results (arising from reporting biases) for each synthesis assessed. | Page 27 |
| Certainty of evidence | 22 | Present assessments of certainty (or confidence) in the body of evidence for each outcome assessed. | n.a. |
| **DISCUSSION** | | |  |
| Discussion | 23a | Provide a general interpretation of the results in the context of other evidence. | Pages 27-32 |
|  | 23b | Discuss any limitations of the evidence included in the review. | Page 32 |
|  | 23c | Discuss any limitations of the review processes used. | Page 32 |
|  | 23d | Discuss implications of the results for practice, policy, and future research. | Page 33 |
| **OTHER INFORMATION** | | |  |
| Registration and protocol | 24a | Provide registration information for the review, including register name and registration number, or state that the review was not registered. | Page 6 |
|  | 24b | Indicate where the review protocol can be accessed, or state that a protocol was not prepared. | Page 6 |
|  | 24c | Describe and explain any amendments to information provided at registration or in the protocol. | n.a. |
| Support | 25 | Describe sources of financial or non-financial support for the review, and the role of the funders or sponsors in the review. | Page 34 |
| Competing interests | 26 | Declare any competing interests of review authors. | Page 34 |
| Availability of data, code and other materials | 27 | Report which of the following are publicly available and where they can be found: template data collection forms; data extracted from included studies; data used for all analyses; analytic code; any other materials used in the review. | n.a. |

**S4 Table. Summary of bias using the Joanna Briggs Checklist for Diagnostic Test Accuracy**

| **Study** | **Total questions assessed** | **Total with indication of bias (answer = No)** | **% questions with indication of bias** | **Bias risk classification** |
| --- | --- | --- | --- | --- |
| Alamo, 2012 | 5 | 2 | 40% | Moderate risk |
| Auld, 2020 | 8 | 2 | 25% | Moderate risk |
| Auld, 2021 | 8 | 2 | 25% | Moderate risk |
| Aunsborg, 2020 | 10 | 1 | 10% | Low risk |
| Awolude, 2021 | 10 | 0 | 0% | Low risk |
| Baik, 2020 | 10 | 2 | 20% | Low risk |
| Balcha, 2014 | 8 | 1 | 13% | Low risk |
| Balkus, 2018 | 8 | 0 | 0% | Low risk |
| Balkus, 2016 | 9 | 0 | 0% | Low risk |
| Brown, 2012 | 9 | 2 | 22% | Moderate risk |
| Hanifa, 2017 | 8 | 2 | 25% | Moderate risk |
| Kahle, 2013 | 8 | 1 | 13% | Low risk |
| Kerschberger, 2021 | 9 | 1 | 11% | Low risk |
| Khan, 2014 | 8 | 0 | 0% | Low risk |
| Maskew, 2020 | 7 | 0 | 0% | Low risk |
| Mbu, 2018 | 10 | 0 | 0% | Low risk |
| Mellins, 2017 | 9 | 1 | 11% | Low risk |
| Mlisana, 2013 | 10 | 1 | 10% | Low risk |
| Modi, 2016 | 9 | 2 | 22% | Moderate risk |
| Njuguna, 2022 | 8 | 1 | 13% | Low risk |
| Peebles, 2020 | 8 | 1 | 13% | Low risk |
| Rosen, 2019 | 8 | 0 | 0% | Low risk |
| Semitala, 2019 | 6 | 2 | 33% | Moderate risk |
| Skogmar, 2014 | 10 | 1 | 10% | Low risk |
| Surka, 2014 | 6 | 3 | 50% | Moderate risk |
| Wahome, 2013 | 9 | 1 | 11% | Low risk |
| Wahome, 2018 | 9 | 1 | 11% | Low risk |
| Wall, 2021 | 8 | 2 | 25% | Moderate risk |
